# Enhancing Healthcare Accessibility: A Comprehensive Usability Study of Pathpoint® eDerma Software

**DOI:** 10.1101/2024.05.17.24307401

**Authors:** Balamurugan Subramaniyan, Atlas Naqvi, Muna Mohamud, Piyush Mahapatra

## Abstract

This research presents a comprehensive usability evaluation of Pathpoint® eDerma Software, conducted in accordance with IEC 62366-1:2015 standards. The study encompasses four diverse user groups, that includes administrators, medical photographers, dermatologists, and patients, ensuring a holistic assessment of the software’s usability and effectiveness. Through a combination of quantitative metrics and qualitative feedback, the study explores various aspects such as accessibility, navigation, safety, and user satisfaction. Potential risks to user experience and patient data security are identified and addressed to ensure compliance with safety standards. The findings highlight the software’s effectiveness in facilitating remote assessments, streamlining workflows, and improving patient care. This research contributes valuable insights to the ongoing refinement and optimisation of eDerma Software, aiming to enhance its usability, safety, and overall effectiveness in real-world healthcare settings.

## 1. Introduction

The dynamic and ever-evolving field of medical device creation places a growing emphasis on the necessity of focusing on usability. This has been underscored by standard 62366, which was issued by the International Electrotechnical Commission (IEC). This standard is a vital guide to integrating usability engineering processes throughout the different stages of medical device design and production [ISO, 2015; Wiklund and Kendler, 2018; IEC, 2016; Karshmer and Tenenberg, 2016].

The field of medical device development recognises the significance of usability studies, as demonstrated by research conducted by Bevan and Macleod in 1994. Their work highlights the necessity of assessing usability by examining productivity, effectiveness and user satisfaction. They also explored different methodologies for evaluating usability, offering a thorough understanding of the subject. The ESPRIT MUSiC project, which provided tools for measuring usability in lab and field settings, is another significant contribution to the field [Virzi, 1992].

Pioneering works by Virzi in 1992, Nielsen in 1993, and Lewis in 1995 have greatly influenced the field of usability studies. They have offered valuable insights into fine-tuning the test phase of usability studies and validated tools for measuring user satisfaction. These works have played a crucial role in guiding the direction of usability.

The development of the System Usability Scale (SUS) by Brooke in 1996 signifies a significant advancement in the field of usability studies. This tool is easy and quick to use, making it a valuable resource for both researchers and developers [Brooke, 1996]. Dumas and Redish’s comprehensive guide to usability testing, published in 1999, is another key resource that offers an in-depth guide to performing usability tests [Dumas and Redish, 1999]. Shneiderman’s influential book on user interface design, published in 1998, is a rich source of knowledge and methodologies in the field [Shneiderman, 1998].

Building on this robust foundation, Kushniruk and Patel’s 2004 work concentrates on cognitive and usability engineering methods in evaluating clinical information systems. Their work offers a thorough understanding of the subject and valuable insights into the field [Kushniruk and Patel, 2004]. The 2006 study by Marcilly, Ammenwerth, and Roehrer examines the usability of user interfaces for clinical information systems and investigates effective evaluation frameworks [Marcilly et al., 2006]. Rubin and Chisnell’s Handbook, published in 2008, offers practical methodologies for executing effective usability tests [Rubin and Chisnell, 2008].

The convergence of usability studies and medical devices is a current research focus. This is evidenced by the work of Patel et al., who conducted usability testing within the framework of IEC 62366 [Patel et al., 2017]. Hignett et al.’s focus on usability engineering in medical device development is another substantial contribution to the field [Hignett et al., 2018]. The exploration of IEC 62366 integration by Nielsen et al. and the review of usability evaluation methodologies and IEC 62366 compliance by Smith et al. underline ongoing efforts to align usability practices with regulatory standards [Nielsen et al., 2019; Smith et al., 2020].

In this context, our research seeks to add to this body of knowledge by providing a thorough usability evaluation and performance review of the Pathpoint® eDerma Software. This study is based on the principles of IEC 62366 and draws on established usability methodologies. This study seeks to improve medical technologies’ overall usability and user experience. We aim to enhance patient outcomes and user satisfaction by conforming to regulatory standards. This research is a significant step towards these goals and contributes to the ongoing efforts to improve the usability of medical devices.

## 2. Overview and Importance of Usability in Pathpoint® eDerma Software

### 2.1 Overview

Pathpoint® eDerma is a software designed to streamline the process of dermatological care by facilitating efficient communication, data management, and remote assessment between medical photographers, dermatologists, and patients. Its primary aim is to enable accurate and timely diagnosis of skin conditions through the seamless capture, storage, and evaluation of derma images, coupled with effective collaboration among healthcare professionals.

The cloud-based, Software-as-a-service (SaaS) model on which the Pathpoint® eDerma Software operates is a revolutionary approach in the healthcare industry. This model allows the software to be accessed from any location and at any time, providing healthcare providers with the flexibility they need to deliver high-quality care. In addition, the SaaS model offers scalability, allowing healthcare providers to expand or reduce their usage based on their needs quickly. This scalability ensures that healthcare providers only pay for what they use, making the software a cost-effective solution.

Another significant advantage of the SaaS model is its accessibility. With the cloud-based software, healthcare providers can access it from any device with an internet connection. This accessibility ensures that healthcare providers can access patient information quickly and easily, enhancing the quality of care provided. Moreover, the SaaS model eliminates the need for healthcare providers to invest in expensive hardware or software, further enhancing its cost-effectiveness.

A vital feature of the Pathpoint® eDerma Software is its seamless integration with electronic healthcare records. This integration ensures smooth and efficient data exchange, eliminating the potential for errors that can occur with manual data entry. By automating data exchange, the software reduces the time and effort healthcare providers require to access and update patient information. This efficiency enhances the quality of care provided and allows healthcare providers to focus more on patient care rather than administrative tasks.

The seamless integration of the Pathpoint® eDerma Software with electronic healthcare records also ensures that patient information is up-to-date and accurate. This accuracy is crucial in healthcare, as it ensures that healthcare providers have the most recent and accurate information when making decisions about patient care. By ensuring the accuracy of patient information, the software enhances the effectiveness of healthcare delivery, leading to improved patient outcomes.

Regarding security, the Pathpoint® eDerma Software aligns with industry-standard security protocols. This alignment ensures that sensitive patient data is protected from potential threats, demonstrating Open Medical’s commitment to data privacy and security. By aligning with industry-standard security protocols, the software ensures that patient data is stored and transmitted securely, reducing the risk of data breaches.

The software’s compliance with NHS Digital standards further validates its reliability and effectiveness. These standards set out stringent requirements for healthcare technology, covering aspects such as interoperability, data quality, and security. By meeting these requirements, the Pathpoint® eDerma Software demonstrates its ability to deliver reliable, high-quality service. The compliance with NHS Digital standards also ensures that the software is compatible with other healthcare technologies, enhancing its interoperability.

In addition to complying with NHS Digital standards, the software adheres to the Web Content Accessibility Guidelines. These guidelines ensure the software is accessible to all users, including those with disabilities. This commitment to accessibility underscores Open Medical’s dedication to making healthcare more inclusive. By adhering to the Web Content Accessibility Guidelines, the software ensures that all users can access and use the software effectively, regardless of their abilities.

Finally, the software has received high ratings for the DTAC criteria by NHS England and ORCHA, a prominent body assessing healthcare technology’s quality and effectiveness. These high ratings further testify to the software’s high performance and reliability. The high ratings from NHS England and ORCHA demonstrate the software’s effectiveness in improving healthcare delivery, further validating its value to healthcare providers.

In summary, the Pathpoint® eDerma Software is a robust, reliable, and effective solution for healthcare providers. Its seamless integration with electronic healthcare records, compliance with stringent industry standards, and commitment to accessibility and security make it an excellent choice for healthcare providers looking to enhance their service delivery. With its innovative features and proven effectiveness, the Pathpoint® eDerma software is set to revolutionise healthcare delivery, improving patient outcomes and enhancing healthcare efficiency.

### 2.2 Intended Use

Pathpoint® eDerma is a cloud-based dermatology platform designed to assist healthcare professionals in documenting and managing dermatological conditions. It is intended to be a fully interoperable application on compatible computing devices with electronic healthcare records (EHRs) and other referral systems, such as e-RS and GP Connect.

Pathpoint® eDerma is intended to be used by qualified healthcare professionals, including dermatologists, dermatology specialty trainees, and other medical practitioners involved in diagnosing and treating skin conditions. The software provides a digital platform for capturing, storing, and analysing patient information related to dermatological examinations, assessments, and treatments.

### 2.3 Environment

Pathpoint® eDerma operates as a teledermatology system designed to facilitate screening patients with skin conditions. The application can be classified into two based on the involvement of the Pathpoint® eDerma Platform. They are

- Active teledermatology and
- Passive teledermatology

#### 2.3.1 Active Teledermatology

Active teledermatology is designed to support patients from their GP or a local Community Diagnostic Centre in rural or distant settings. This type is termed “active,” as patients must visit their local facility for initial diagnostics. The patient pathway is outlined as follows: individuals with concerning skin symptoms holding prospective appointments at community health centres respond to a teledermatology questionnaire they receive in their email via the Pathpoint® eDerma platform before attending the community health centre.

At the health centre, a medical photographer (photographer) or healthcare worker equipped with a DSLR camera or a smartphone with a dermatoscope attachment captures images of the area(s) of concern on the patient’s skin. Subsequently, these images are uploaded to the Pathpoint® eDerma platform, where they are available to a consultant dermatologist who reviews the patients’ questionnaire responses and photographs of the skin lesions. Based on this review, the dermatologist plans further treatment, including a diagnosis plan for suspected cancer or appropriate measures for non-concerning cases. This pathway is termed “active” due to the active data collection directly from the patient (in the form of the Teledermatology Questionnaire and images).

The block diagram of the flow in active teledermatology is shown in Figure 1.

**Figure 1:**
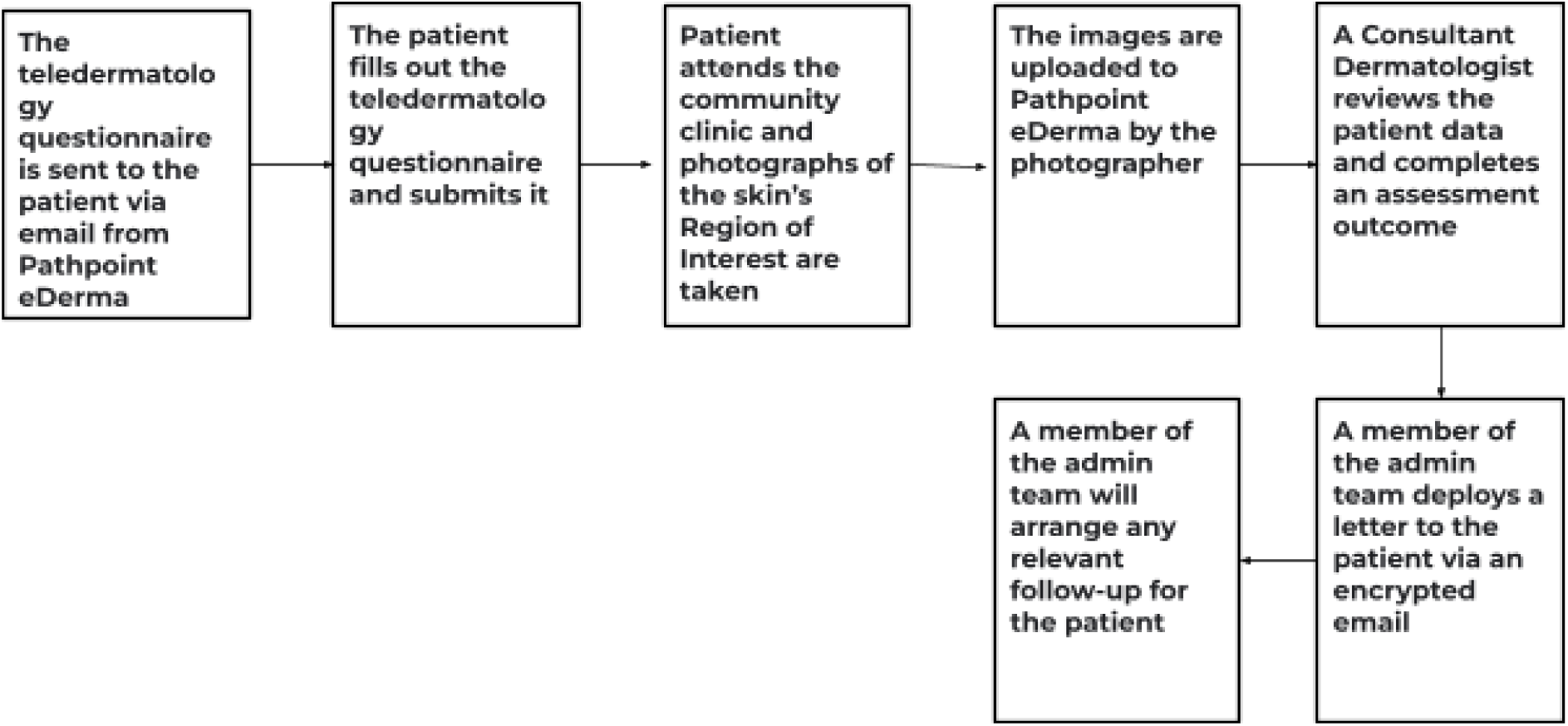
Active Teledermatology

#### 2.3.2. Passive Teledermatology

Passive teledermatology, on the other hand, only involves the patient interacting with eDerma at the end of their diagnostic journey. Instead, patient data is consolidated from other patient data systems, such as the Electronic Referral Service (eRS). Pathpoint® eDerma, in this scenario, only retrieves and summarises this data for the dermatologist. The patient is not required to fill out a questionnaire or attend a clinic specifically for photography. For this reason, the critical differentiator between the active and passive pathways is the level of involvement required from the patients.

The block diagram of the patient flow in passive teledermatology is shown in Figure 2.

**Figure 2:**
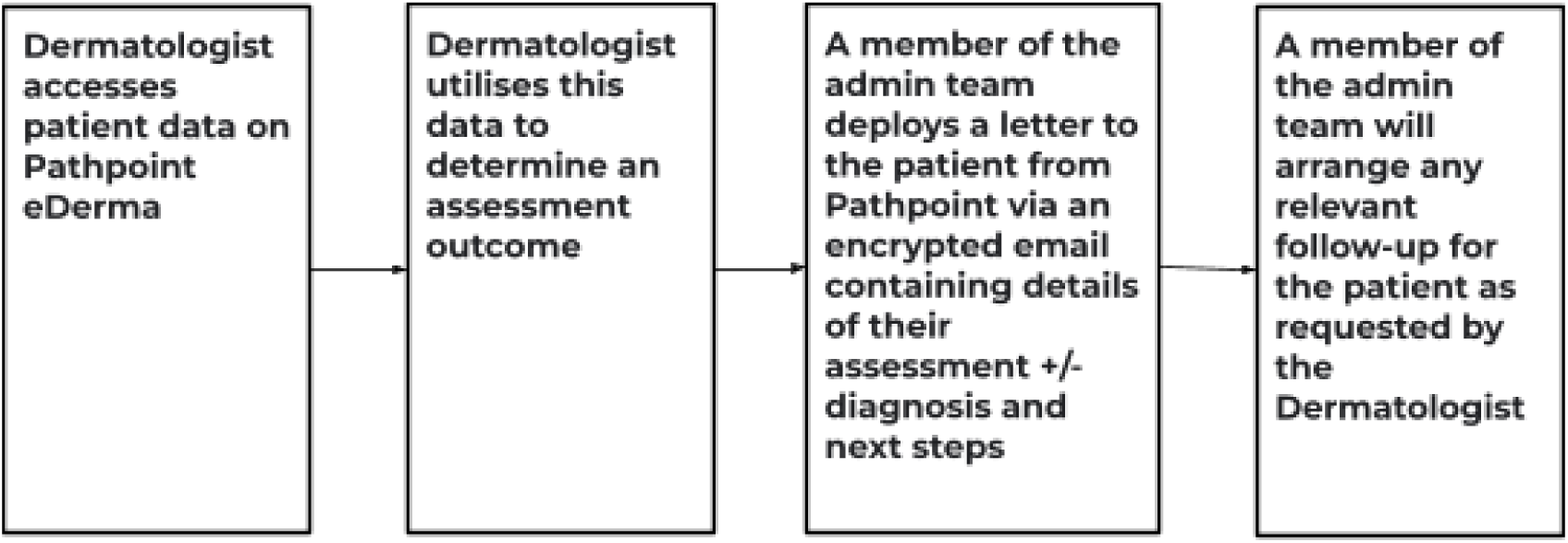
Passive Teledermatology.

### 2.3 Why Usability is Critical for Pathpoint® eDerma

Usability plays a crucial role in the effectiveness and adoption of Pathpoint® eDerma for several reasons:

1. **Efficiency:** Streamlined workflows and intuitive interface design ensure that healthcare professionals can perform tasks quickly and accurately, reducing administrative burden and saving time.
2. **Accuracy:** Clear navigation and user-friendly features promote accurate data entry and interpretation, minimising errors and enhancing the reliability of diagnoses and treatment plans.
3. **Accessibility:** User-centric design considerations cater to the diverse needs of healthcare professionals and patients, ensuring that the software is accessible to individuals with varying levels of technical expertise and physical abilities.
4. **Patient Engagement:** Intuitive patient-facing features enhance engagement and compliance by making it easy for patients to provide information, access their records, and participate in remote assessments, ultimately improving health outcomes.
5. **Adoption:** Positive user experiences drive user acceptance and adoption of the software, leading to increased usage and improved collaboration among healthcare professionals, which in turn benefits patient care and overall healthcare delivery.

In summary, ensuring the usability of Pathpoint® eDerma is paramount for maximising its potential to streamline dermatological care, improve diagnostic accuracy, enhance patient engagement, and, ultimately, positively impact patient outcomes.

## 3. Methods

The usability procedures and documentation of the software were diligently aligned with the requirements of IEC 62366-1:2015 to guarantee compliance with the set standards. This action was vital as it confirmed that the software was developed in accordance with the globally accepted standards for usability. The IEC 62366-1:2015 standard is an exhaustive guideline that provides a structure for implementing usability engineering in medical devices. It details the essential steps and protocols to ensure the software is user-friendly, safe, and effective for its target audience.

Essential sections, such as Information for Safety related to Usability and the Usability Engineering File, were vital to the assessment process. These sections offered critical information needed for the all-inclusive assessment of the software’s usability. The Information for Safety related to Usability section is fundamental to the IEC 62366-1:2015 standard. It gives users the necessary information concerning the software’s safe and effective utilisation. This section comprises detailed information about possible hazards, warnings, and precautions that users must be aware of when operating the software. It is designed to ensure users have all the required information to use the software safely and effectively.

The Usability Engineering File is a complete document that comprises all relevant information about the software’s usability. It includes detailed documentation of the usability engineering procedures, methodologies, and results that have been documented throughout the software’s development lifecycle. This file functions as a comprehensive record of the software’s usability, offering a detailed overview of the software’s usability attributes and functionalities.

Usability activities followed the risk management procedure, guided by vital documents such as the Hazard-Related Use Scenarios list, Usability Evaluation Protocol, and Usability Evaluation Plan. This strategy ensured that usability activities were systematically conducted and controlled, guaranteeing the overall safety and effectiveness of the software.

The risk management procedure is a systematic approach to identifying, evaluating, and mitigating usability risks. It involves an in-depth analysis of the software’s attributes and functionalities, identifying potential usability risks, evaluating the severity of these risks, and implementing suitable measures to mitigate these risks. This strategy ensures that potential harm to users is minimised and that the overall safety of the software is guaranteed.

Several critical documents were used to support the risk management process. One such document was the Hazard-Related Use Scenarios list. This list identifies potential hazards users may encounter while operating the software and describes the corresponding use scenarios. By identifying and understanding these hazards, suitable usability measures can be implemented to mitigate the associated risks.

The Usability Evaluation Protocol was another vital document used in the risk management process. This protocol outlines the specific procedures and methods for conducting usability evaluations. It ensures consistency and repeatability in the evaluation process, ensuring the software’s usability is evaluated consistently and reliably.

The Usability Evaluation Plan provides a comprehensive overview of the entire usability evaluation process. This plan includes details about the objectives, scope, methods, participants, and evaluation schedule. It functions as a roadmap for conducting the evaluations, ensuring that all necessary aspects of the usability evaluation process are covered.

By adhering to these established standards, conducting usability activities in line with a risk management procedure, and using vital documents such as the Hazard-Related Use Scenarios list, Usability Evaluation Protocol, and Usability Evaluation Plan, the software’s usability was thoroughly evaluated and guaranteed to meet the necessary standards for safety and effectiveness.

This comprehensive approach to usability evaluation guarantees that the software is user-friendly, safe and effective for its target audience. It guarantees that the software meets the highest usability standards, offering users a dependable, safe, and effective tool for their needs. This approach to usability evaluation is stringent and methodical, ensuring that each facet of the software’s usability is thoroughly examined and adjusted to the highest usability standards.

## 4. User Group

The study comprises four distinct user groups:

- Patients,
- Dermatologists,
- Administrators, and
- Medical Photographers.

The first three user groups (Patients, dermatologists, and administrators) are common to active and passive teledermatology, as their interactions with the software remain largely consistent across both modes. The photographers’ group is a specific user group pertaining only to active teledermatology.

### 4.1 Patients

In active teledermatology, patients participate by responding to a teledermatology questionnaire related to their skin condition and attending a photography appointment, providing essential information about their skin condition. Patients receive email correspondence in active and passive teledermatology regarding their assessment and/or outcome.

### 4.2 Dermatologists

Dermatologists are tasked with reviewing patient referral information, questionnaire responses, and skin lesion images in the case of active teledermatology. In passive teledermatology, they review data from other eRS solutions that have been added either manually or via integration to Pathpoint®. Based on this information, dermatologists make informed decisions regarding the patient’s possible diagnosis and treatment. Following the review, dermatologists can communicate outcomes to patients via email and, if necessary, refer them for follow-up or procedures.

### 4.3 Administrators

Administrators (admins) are crucial in ensuring the smooth operation of the Pathpoint® eDerma platform. Administrative teams will usually have an overview of the patient journey through eDerma. Specific administrative tasks include reviewing lists, actioning outcomes, and ensuring HCP tasks are also completed in a timely manner.

### 4.4 Medical Photographers (Healthcare Assistants)

Health workers, including medical photographers and healthcare assistants in active settings, are crucial in capturing images of patients’ skin lesions. Their primary responsibility is to take photographs and expertly upload them to the eDerma platform. Proficiency in photography (often including dermatoscopy) is expected.

## 5. Tasks List

The task list differs for the user groups and is carefully designed based on the hazardous use scenarios after proper risk analysis. They have been individually explained below based on their interactions with the platform and the impact on the usability element of the software platform. The outcomes of the usability study will be presented in the usability report.

The selection of tasks for the usability evaluation was meticulously crafted to align with the distinct roles and expectations of the four user groups. Each task was thoughtfully chosen to assess the critical functionalities of the Pathpoint® eDerma Software relevant to the respective user group’s interactions with the platform. The rationale behind the task selection is outlined below:

### User-Centric Approach

Tasks for the patients’ user group were designed to focus on their primary interaction with the Pathpoint® eDerma Questionnaire. This includes clicking the questionnaire link, reading and answering questions, and submitting responses. The goal was to evaluate the platform’s accessibility and user-friendliness for individuals with varying demographics and technological proficiencies.

For the other user groups, which are health professionals (medical photographers, dermatologists, and administrations), tasks were tailored to assess their interaction with the software across various responsibilities. This included login procedures, patient data management, and additional functionalities specific to healthcare professionals, such as sending reminders and interpreting patient data. The aim was to ensure that the software aligns seamlessly with the workflows and needs of healthcare professionals, contributing to optimal efficiency in clinical settings.

### Comprehensive System Evaluation

Tasks for all the user groups covered a range of functionalities, from fundamental interactions like accessing the questionnaire to more complex actions such as generating and deploying questionnaires, interpreting patient data, and utilising support features. This comprehensive approach aimed to provide a holistic evaluation of the Pathpoint® eDerma Software’s usability, ensuring that it meets the diverse needs of its user base.

### Risk Mitigation

Tasks were selected to cover critical functionalities where failure could lead to potential use errors or hazards. For instance, tasks involving sending reminders, updating patient details, and interpreting patient data were included to assess the software’s robustness in managing healthcare-related information and communication.

### Diversity and Inclusivity

The task list considered the user groups’ diverse backgrounds and capabilities. For the patient population user group, tasks were designed to accommodate users with varying technological proficiencies. For the other user groups, tasks addressed the specific needs of healthcare professionals, acknowledging their advanced training and responsibilities.

### End-to-End Usability

The inclusion of tasks that spanned the entire user journey, from login procedures to end-to-end questionnaire completion, aimed to evaluate the software’s usability across different stages. This approach helps ensure a positive and streamlined user experience throughout the process.

The task list differs for all four user groups. They have been individually tabulated below, and the results will be presented in the usability report.

### 5.1 Patients

The task list for patients is meticulously designed to ensure a seamless and user-friendly experience while interacting with the eDerma platform. The primary objective is to gather essential information about the patient’s skin condition through a teledermatology questionnaire for patients in the active teledermatology pathway. In active and passive teledermatology scenarios, patients share pertinent details regarding their skin lesions by responding to the questionnaire. The overview of the task to be performed is below. A detailed list can be found in Table 1.

**Table 1:**
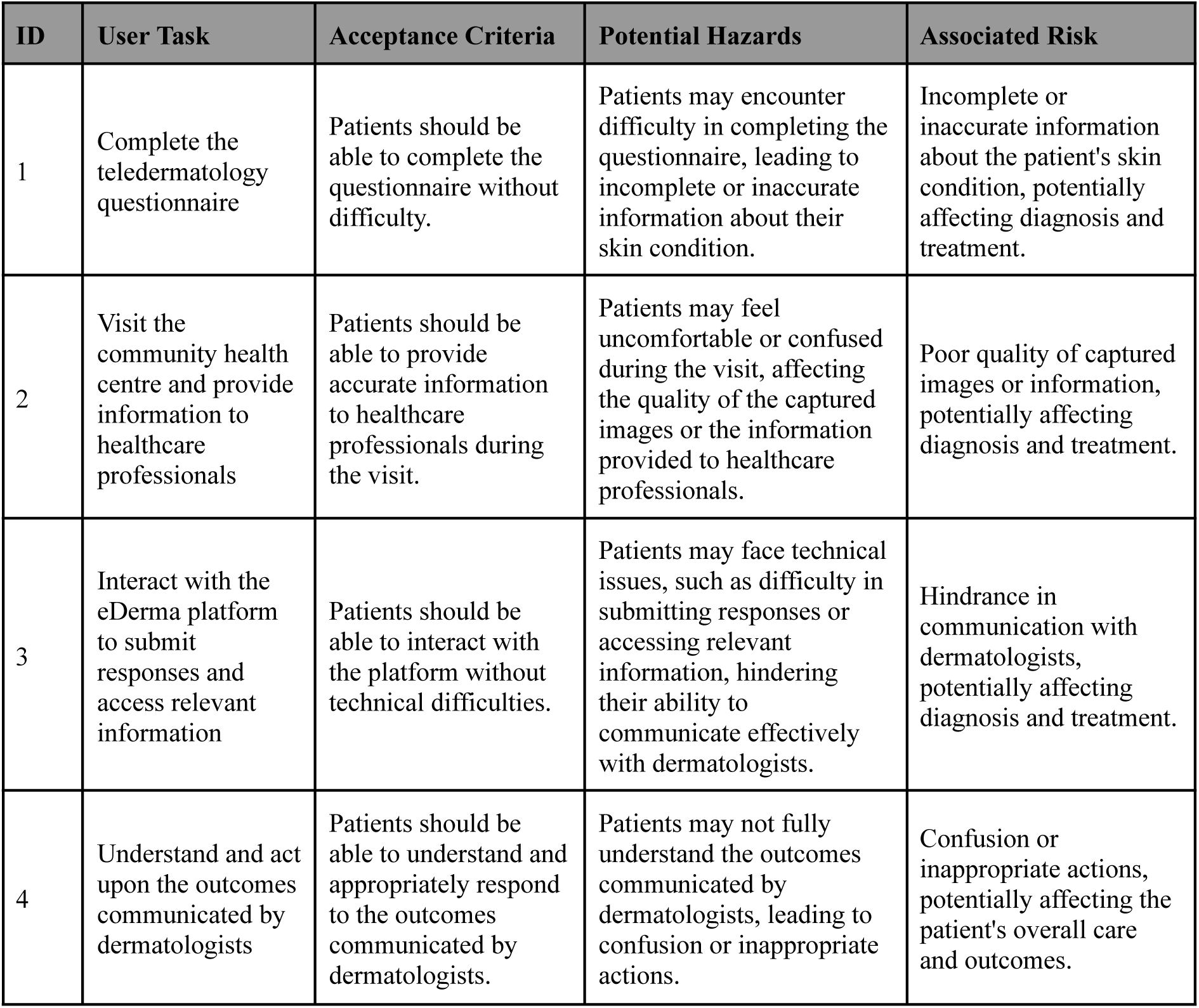
User Group: Patients.

| ID | User Task | Acceptance Criteria | Potential Hazards | Associated Risk |
| --- | --- | --- | --- | --- |
| 1 | Complete the teledermatology questionnaire | Patients should be able to complete the questionnaire without difficulty. | Patients may encounter difficulty in completing the questionnaire, leading to incomplete or inaccurate information about their skin condition. | Incomplete or inaccurate information about the patient's skin condition, potentially affecting diagnosis and treatment. |
| 2 | Visit the community health centre and provide information to healthcare professionals | Patients should be able to provide accurate information to healthcare professionals during the visit. | Patients may feel uncomfortable or confused during the visit, affecting the quality of the captured images or the information provided to healthcare professionals. | Poor quality of captured images or information, potentially affecting diagnosis and treatment. |
| 3 | Interact with the eDerma platform to submit responses and access relevant information | Patients should be able to interact with the platform without technical difficulties. | Patients may face technical issues, such as difficulty in submitting responses or accessing relevant information, hindering their ability to communicate effectively with dermatologists. | Hindrance in communication with dermatologists, potentially affecting diagnosis and treatment. |
| 4 | Understand and act upon the outcomes communicated by dermatologists | Patients should be able to understand and appropriately respond to the outcomes communicated by dermatologists. | Patients may not fully understand the outcomes communicated by dermatologists, leading to confusion or inappropriate actions. | Confusion or inappropriate actions, potentially affecting the patient's overall care and outcomes. |

#### 5.1.1. Teledermatology Questionnaire

- **Objective:** Patients in the active teledermatologic pathway are tasked with completing a comprehensive teledermatology questionnaire to provide crucial information about their skin condition.
- **Scenario:** In both active teledermatology, patients engage with the questionnaire, detailing symptoms, medical history, and other relevant information.

#### 5.1.2. Community Health Centre Visit (Active Teledermatology Only)

- **Objective**: In the active teledermatology setting, patients are required to visit the community health centre, where an appropriately trained healthcare professional will capture photographs of their skin lesion(s).
- **Scenario:** Patients attend their GP practice or a local Community Diagnostic Centre as part of the active data collection process. This allows a designated healthcare professional to capture images of their skin lesions.

#### 5.1.3. Interacting with eDerma Platform

- **Objective:** Patients navigate and interact with the eDerma platform to submit questionnaire responses and access relevant information.
- **Scenario:** Patients use the platform to seamlessly provide responses to the teledermatology questionnaire, fostering a user-friendly interaction.

#### 5.1.4. Reviewing Outcome

- **Objective:** Patients receive and review outcomes communicated by dermatologists via an encrypted email.
- **Scenario:** After the dermatologist’s review, patients are informed about their possible diagnosis and proposed next steps, including, if needed, referrals for further diagnosis or treatment.

This patient-centric task list aims to ensure that individuals with suspected skin diseases actively engage with the platform, fostering effective communication with healthcare professionals and contributing valuable information for the diagnostic process.

### 5.2 Dermatologist

The task list for dermatologists is strategically crafted to leverage their expertise in diagnosing and managing skin conditions through the eDerma platform. The tasks are designed to facilitate a comprehensive review of patient data, enabling informed decision-making and effective communication with patients as well as the administrative teams. The overview of the task to be performed is below. A detailed list can be found in Table 2.

**Table 2:** User Group: Dermatologist.

| ID | User Task | Acceptance Criteria | Potential Hazards | Associated Risk |
| --- | --- | --- | --- | --- |
| 1 | Reviewing Patient Questionnaire Responses (Active Teledermatology) | Dermatologists should be able to thoroughly examine the responses provided by patients in the teledermatology questionnaire without difficulty. | Dermatologists may not be able to understand the priority of the patients based on their list, leading to delays in diagnosing and treating patients with urgent or critical conditions, potentially compromising patient outcomes and safety. | Delayed diagnosis and treatment of urgent or critical conditions, potentially compromising patient outcomes and safety. |
| 2 | Evaluating Skin Lesion Images (Active Teledermatology) | Dermatologists should be able to scrutinise images of skin lesions captured by photographers in community health centres without difficulty. | Inconsistent navigation may impede workflow efficiency and increase the cognitive load on dermatologists, leading to errors or missed opportunities for accessing critical patient data. | Impaired workflow efficiency, increased cognitive load, errors, and missed opportunities for accessing critical patient data. |
| 3 | Utilising Data from Other eRS Solutions (Active & Passive Teledermatology) | Dermatologists should be able to leverage data retrieved from other eRS solutions, such as clinical history and referral information, without difficulty. | Inadequate information display may result in dermatologists overlooking critical information or misinterpreting patient data, leading to diagnosis or treatment planning errors. | Overlooking critical information and misinterpreting patient data, resulting in diagnosis or treatment planning errors. |
| 4 | Decision-Making for Prognosis | Dermatologists should be able to make informed decisions regarding the prognosis of patients based on the reviewed data without difficulty. | Insufficient training and support may lead to dermatologists struggling to use the eDerma platform effectively, resulting in reduced confidence in its capabilities and reluctance to incorporate it into their clinical practice. | Reduced confidence in the platform, reluctance to incorporate it into clinical practice, workflow disruptions, and missed opportunities for leveraging its full potential in patient care. |
| 5 | Communicating Outcomes to Patients | Dermatologists should be able to convey diagnostic outcomes and recommended actions to patients via an encrypted email without difficulty. | Not being able to interpret the flow in Pathpoint may lead to errors in data entry, missed steps in the diagnostic process, or frustration among users. | Errors in data entry, missed steps in the diagnostic process, and frustration among users. |
| 6 | Accessing Patient Information | Dermatologists should be able to access up-to-date patient information without difficulty. | Incomplete integration with EHR systems may result in difficulties accessing up-to-date patient information, leading to potential diagnosis or treatment planning errors. | Difficulty accessing up-to-date patient information, diagnosis or treatment planning errors, inefficiencies, detracting from valuable time spent on patient care. |
| 7 | Processing Patient Data | Dermatologists should be able to process patient data effectively without cognitive overload. | Cognitive overload when processing data may impair decision-making and task performance, increasing the likelihood of errors or oversight in patient care. | Impaired decision-making and task performance increased the likelihood of errors or oversight in patient care. |
| 8 | Handling Errors | Dermatologists should be able to diagnose and address technical issues promptly and effectively. | Inadequate error handling may hinder dermatologists' ability to diagnose and address technical issues promptly, leading to frustration, decreased productivity, and potential errors in patient care. | Hindered ability to diagnose and address technical issues promptly, frustration, decreased productivity, potential errors in patient care, trial-and-error approaches, further complications or system instability. |
| 9 | Saving Assessment Information | Dermatologists should be able to save assessment information without difficulty. | Not clicking the Save Assessment button may result in difficulties in saving the assessment information, making onward actions not possible. | Loss of assessment information, inability to proceed with onward actions. |
| 10 | Applying Filters | Dermatologists should be able to apply filters correctly to select the appropriate patient list. | Incorrect filter application can lead to the wrong list of patients being actioned, resulting in delayed diagnosis for those pending on the correct list | Delayed diagnosis |
| 11 | Filtering Referrals | Dermatologists should be able to filter referrals effectively. | Difficulty in filtering referrals may result in missed or delayed access to relevant patient information, hindering timely diagnosis and treatment. | Missed or delayed access to relevant patient information, hindering timely diagnosis and treatment. |
| 12 | Evaluating Skin Lesions | Dermatologists should be able to evaluate skin lesions accurately. | Wrong evaluation of the skin lesion may compromise the accuracy of dermatologists' diagnoses, leading to inadequate patient management or unnecessary interventions. | Inadequate patient management and unnecessary interventions. |
| 13 | Managing Workflow | Dermatologists should be able to manage their workflow efficiently. | Multiclick issue may slow down workflow processes, decrease user productivity, and contribute to user dissatisfaction with the platform. | Slowed workflow processes, decreased user productivity, and user dissatisfaction. |
| 14 | Using SNOMED CT | Dermatologists should be able to use SNOMED CT comfortably and proficiently. | Discomfort with SNOMED CT may lead to errors, incomplete or inaccurate documentation, and potential difficulties in interoperability with other healthcare systems. | Errors, incomplete or inaccurate documentation, and difficulties in interoperability with other healthcare systems. |
| 15 | Reviewing Images | Dermatologists should be able to review images accurately. | The wrong image was uploaded for review may result in misdiagnosis, inappropriate treatment decisions, or delays in patient care. | Misdiagnosis, inappropriate treatment decisions or delays in patient care. |

#### 5.2.1. Reviewing Patient Questionnaire Responses (Active Teledermatology)

- **Objective:** Dermatologists thoroughly examine the responses provided by patients in the teledermatology questionnaire.
- **Scenario:** Dermatologists analyse the questionnaire to gather information about symptoms, medical history, and other relevant details, forming the basis for the diagnostic process.

#### 5.2.2. Evaluating Skin Lesion Images (Active Teledermatology)

- **Objective:** In active teledermatology, dermatologists scrutinise images of skin lesions photographers capture in community health centres.
- **Scenario:** Dermatologists assess the visual data to aid in the diagnosis and determine the appropriate course of action for patients.

#### 5.2.3. Utilising Data from Other eRS Solutions (Active & Passive Teledermatology)

- **Objective:** In both teledermatology pathways, dermatologists leverage data retrieved from other eRS solutions, for example, clinical history and referral information.
- **Scenario**: Dermatologists analyse patient data consolidated from external eRS systems, considering a broader context for diagnosis and treatment planning.

#### 5.2.4. Decision-Making for Prognosis

- **Objective:** Dermatologists make informed decisions regarding the prognosis of patients based on the reviewed data.
- **Scenario:** Taking into account questionnaire responses, skin lesion images, or data from other eRS solutions, dermatologists formulate a potential diagnosis and determine the appropriate next steps for patient management.

#### 5.2.5. Communicating Outcomes to Patients

- **Objective:** Dermatologists convey diagnostic outcomes and recommended actions to patients via an encrypted email.
- **Scenario:** Patients receive clear and comprehensible communications from dermatologists, including information about their diagnosis, potential treatments, and referrals for further diagnosis if necessary.

This task list empowers dermatologists with the tools and information needed to provide accurate diagnoses and treatment plans.

### 5.3 Administrators

The task list for admins is integral to ensuring the smooth operation and coordination of the eDerma platform. Admins play a pivotal role in managing various aspects of patient flowto facilitate efficient teledermatology services. The overview of the task to be performed is below. A detailed list can be found in Table 3.

**Table 3:**
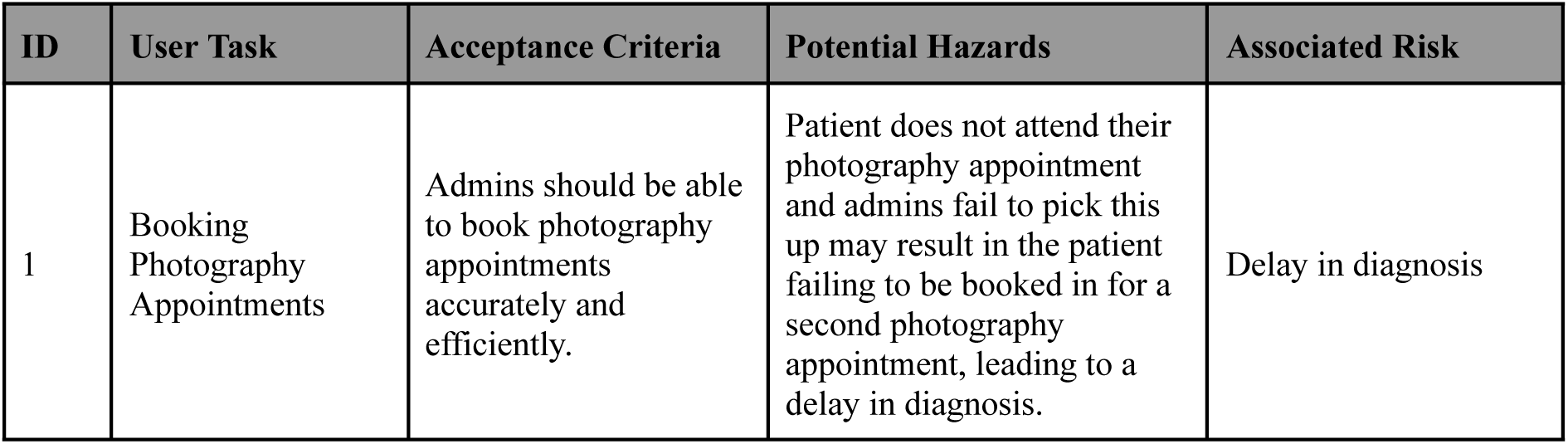
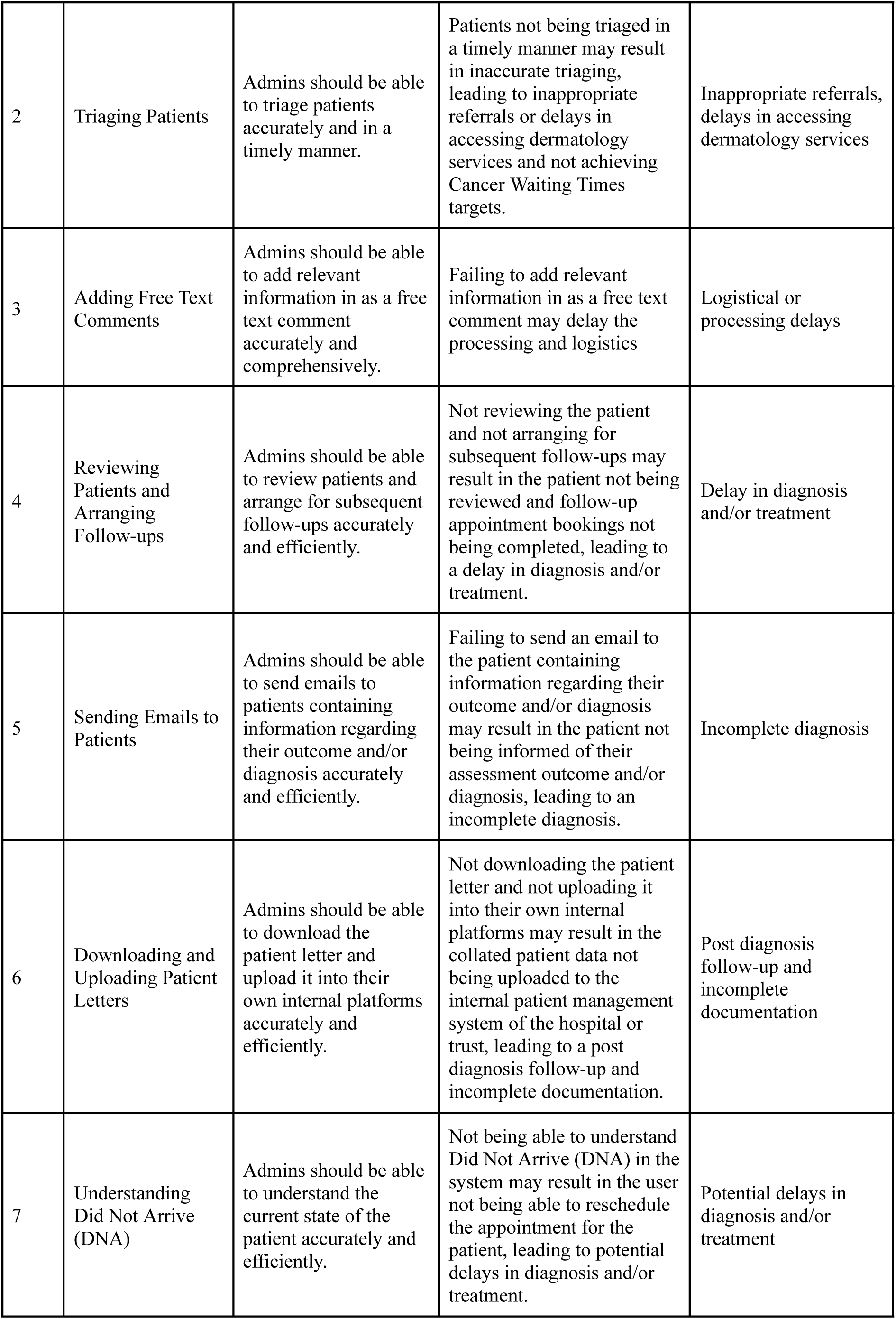
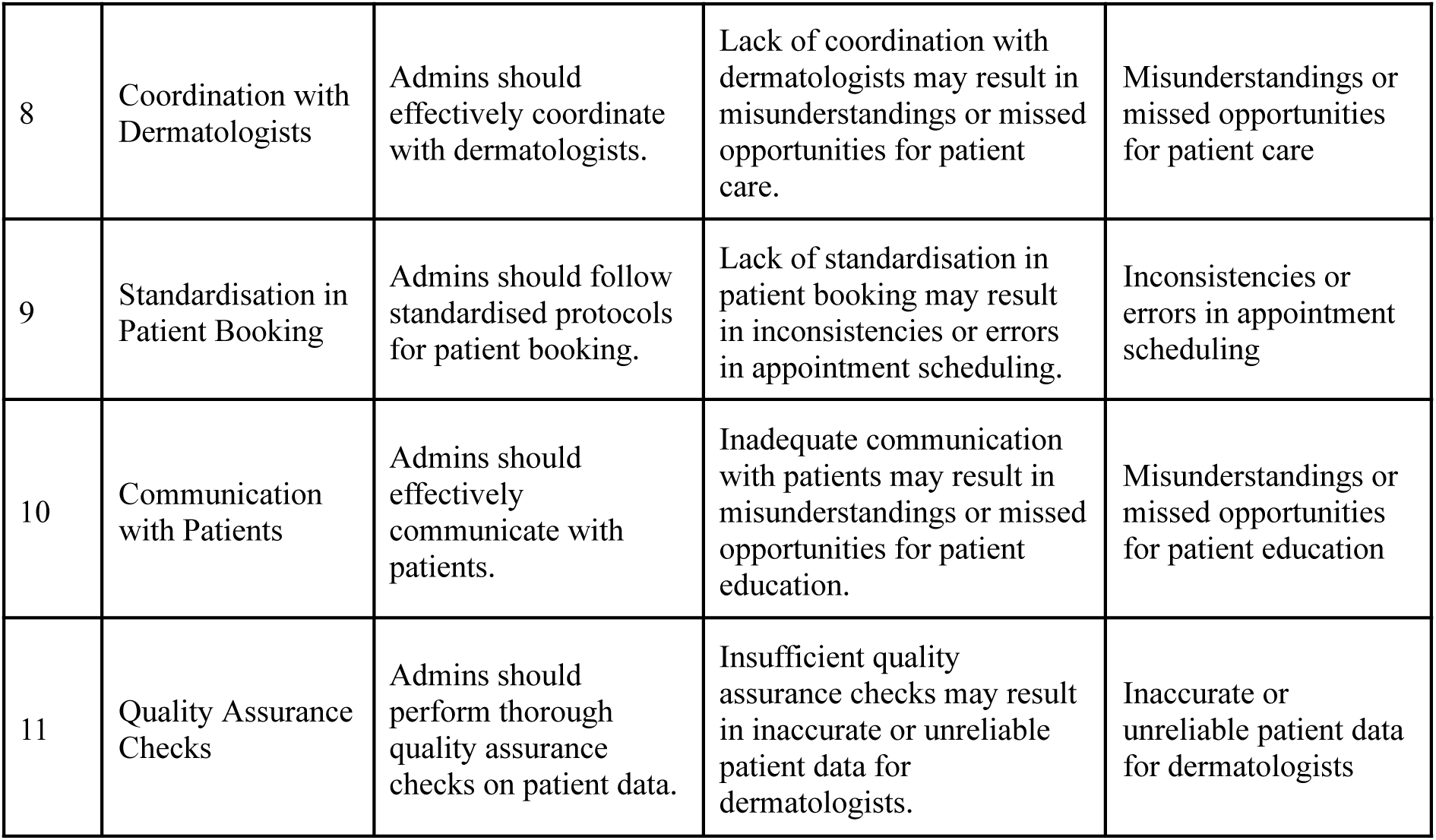
User Group: Administrators.

| ID | User Task | Acceptance Criteria | Potential Hazards | Associated Risk |
| --- | --- | --- | --- | --- |
| 1 | Booking Photography Appointments | Admins should be able to book photography appointments accurately and efficiently. | Patient does not attend their photography appointment and admins fail to pick this up may result in the patient failing to be booked in for a second photography appointment, leading to a delay in diagnosis. | Delay in diagnosis |
| 2 | Triaging Patients | Admins should be able to triage patients accurately and in a timely manner. | Patients not being triaged in a timely manner may result in inaccurate triaging, leading to inappropriate referrals or delays in accessing dermatology services and not achieving Cancer Waiting Times targets. | Inappropriate referrals, delays in accessing dermatology services |
| 3 | Adding Free Text Comments | Admins should be able to add relevant information in as a free text comment accurately and comprehensively. | Failing to add relevant information in as a free text comment may delay the processing and logistics | Logistical or processing delays |
| 4 | Reviewing Patients and Arranging Follow-ups | Admins should be able to review patients and arrange for subsequent follow-ups accurately and efficiently. | Not reviewing the patient and not arranging for subsequent follow-ups may result in the patient not being reviewed and follow-up appointment bookings not being completed, leading to a delay in diagnosis and/or treatment. | Delay in diagnosis and/or treatment |
| 5 | Sending Emails to Patients | Admins should be able to send emails to patients containing information regarding their outcome and/or diagnosis accurately and efficiently. | Failing to send an email to the patient containing information regarding their outcome and/or diagnosis may result in the patient not being informed of their assessment outcome and/or diagnosis, leading to an incomplete diagnosis. | Incomplete diagnosis |
| 6 | Downloading and Uploading Patient Letters | Admins should be able to download the patient letter and upload it into their own internal platforms accurately and efficiently. | Not downloading the patient letter and not uploading it into their own internal platforms may result in the collated patient data not being uploaded to the internal patient management system of the hospital or trust, leading to a post diagnosis follow-up and incomplete documentation. | Post diagnosis follow-up and incomplete documentation |
| 7 | Understanding Did Not Arrive (DNA) | Admins should be able to understand the current state of the patient accurately and efficiently. | Not being able to understand Did Not Arrive (DNA) in the system may result in the user not being able to reschedule the appointment for the patient, leading to potential delays in diagnosis and/or treatment. | Potential delays in diagnosis and/or treatment |
| 8 | Coordination with Dermatologists | Admins should effectively coordinate with dermatologists. | Lack of coordination with dermatologists may result in misunderstandings or missed opportunities for patient care. | Misunderstandings or missed opportunities for patient care |
| 9 | Standardisation in Patient Booking | Admins should follow standardised protocols for patient booking. | Lack of standardisation in patient booking may result in inconsistencies or errors in appointment scheduling. | Inconsistencies or errors in appointment scheduling |
| 10 | Communication with Patients | Admins should effectively communicate with patients. | Inadequate communication with patients may result in misunderstandings or missed opportunities for patient education. | Misunderstandings or missed opportunities for patient education |
| 11 | Quality Assurance Checks | Admins should perform thorough quality assurance checks on patient data. | Insufficient quality assurance checks may result in inaccurate or unreliable patient data for dermatologists. | Inaccurate or unreliable patient data for dermatologists |

#### 5.3.1. Automatic Pulling or Triage

**Objective:** Admins are responsible for automatically pulling or triaging patient data from the Electronic Referral Service (ERS) to determine their suitability for eDerma consultation.

**Scenario:** Admins leverage the integration with ERS to identify and redirect patients to the eDerma platform for further assessment based on predefined criteria such as referral type and urgency.

#### 5.3.2. Patient Booking and Coordination

**Objective:** Admins handle patient booking and coordination for active teledermatology consultations.

**Scenario:** Admins manage the scheduling and coordination of patient appointments, ensuring timely access to dermatology services and effective utilisation of resources.

#### 5.3.3. Downloading and Uploading Documents

**Objective:** Admins download documents from the eDerma platform and upload them into the Electronic Health Record (EHR) system for seamless integration and continuity of patient care.

**Scenario:** Admins retrieve relevant medical documents, reports, and images from eDerma and securely transfer them to the EHR system, ensuring comprehensive patient records and facilitating informed decision-making by healthcare professionals.

#### 5.3.4. Coordinating Patient Pathway

**Objective:** Admins oversee all components of the patient pathway, including follow-up, arrangement, and triage, to ensure seamless coordination and continuity of care.

**Scenario:** Admins actively coordinate patient referrals and follow-up.

The task list for admins is meticulously designed to ensure the smooth setup and operation of the eDerma platform. Admins play a critical role in configuring the eDerma system, managing user accounts, and customising the platform to meet the specific needs of healthcare professionals. The overview of the task to be performed is below. A detailed list can be found in Table 3.

This task list empowers admins with the tools and responsibilities needed to establish and maintain the eDerma platform, ensuring its reliability, security, and effectiveness in supporting teledermatology services.

### 5.4 Medical Photographers

The task list for photographers, who are integral health workers in the active teledermatology setting, is crafted to ensure accurate and detailed capture of images of patients’ skin lesions. Their role involves expertly taking photographs and uploading them to the eDerma platform. The overview of the task to be performed is below. A detailed list can be found in Table 4.

**Table 4:** User Group: Medical Photographers.

| ID | User Task | Acceptance Criteria | Hazards Encountered | Associated Risk |
| --- | --- | --- | --- | --- |
| 1 | Capturing High-Quality Images | Photographers should be able to capture high-quality images of skin lesions accurately and efficiently. | Uploading the derma image(s) in the wrong section may result in misdiagnosis or no diagnosis due to the confusion created. | Misdiagnosis or no diagnosis |
| 2 | Uploading Images to the Platform | Photographers should be able to upload images to the platform accurately and efficiently. | Difficulty in filtering referrals may result in missed or delayed access to relevant patient information, hindering timely diagnosis and treatment. | Missed or delayed access to relevant patient information |
| 3 | Quality Assurance Checks | Photographers should be able to perform quality assurance checks on captured images accurately and efficiently. | Document merger feature failure may result in misdiagnosis or no diagnosis due to the failure in merging the documents. | Misdiagnosis or no diagnosis |
| 4 | Reviewing Patient's Responses to Questionnaire | Photographers should be able to review patient's responses to questionnaires accurately and efficiently. | Failing to review the patient's response to their questionnaire may result in non-diagnosis or misdiagnosis. | Non-diagnosis or misdiagnosis |
| 5 | Accessing QR Code in Software Application | Photographers should be able to access QR code in the software application accurately and efficiently. | Not being able to access QR code in the software application may result in no concrete diagnosis for the patient. | No concrete diagnosis |
| 6 | Uploading Images from Mobile Device | Photographers should be able to upload images from mobile devices accurately and efficiently. | Uploading the wrong image from a mobile device may result in misdiagnosis or no diagnosis due to the confusion created. | Misdiagnosis or no diagnosis |
| 7 | Uploading Derma Pictures | Photographers should be able to upload derma pictures accurately and efficiently. | Not uploading the derma pictures taken may result in no concrete diagnosis for the patient. | No concrete diagnosis |
| 8 | Ensuring Image Quality | Photographers should ensure that the uploaded images are of high quality. | Poor image quality may result in incorrect diagnosis or delayed diagnosis due to insufficient image quality. | Incorrect diagnosis or delayed diagnosis |
| 9 | Collaboration with Dermatologists | Photographers should communicate effectively with dermatologists. | Insufficient collaboration with dermatologists may result in inaccurate diagnosis or delayed diagnosis due to lack of collaboration. | Inaccurate diagnosis or delayed diagnosis |
| 10 | Quality Assurance Checks | Photographers should perform thorough quality assurance checks on captured images. | Inadequate quality assurance checks may result in inaccurate diagnosis or delayed diagnosis due to unreliable image quality. | Inaccurate diagnosis or delayed diagnosis |

#### 5.4.1. Skin Lesion Photography

- **Objective:** As the photographers, they are responsible for capturing high-quality images of patients’ skin lesions.
- **Scenario:** Equipped with a DSLR camera or dermatoscopy with a smartphone, photographers employ their expertise to capture clear and detailed photographs during patients’ visits to the community health centre.

#### 5.4.2. Uploading Images to eDerma Platform

- **Objective:** Photographers upload the captured skin lesion images to the eDerma platform for further analysis by dermatologists.
- **Scenario:** After photographing the skin lesions, photographers use the eDerma platform to upload the images, ensuring timely and secure access for dermatologists.

#### 5.4.3. Expertise in Photography and EPR Platforms

- **Objective:** photographers are expected to possess expertise in photography techniques.
- **Scenario:** Proficiency in capturing quality images and familiarity with the eDerma platform’s functionalities are essential for photographers to contribute effectively to the teledermatology workflow.

#### 5.4.4. Collaboration with Dermatologists

- **Objective:** Photographers collaborate with dermatologists to ensure that captured images provide valuable information for diagnosis.
- **Scenario:** Regular communication with dermatologists may be necessary to address specific requirements or nuances related to the captured images and enhance the overall diagnostic process.

#### 5.4.5. Quality Assurance of Captured Images

- **Objective:** Photographers perform a quality check to ensure that captured images meet the required standards for diagnostic accuracy.
- **Scenario:** This involves assessing factors such as focus, lighting, and clarity to enhance the reliability of the visual data provided to dermatologists.

This task list emphasises the crucial role of Photographers in actively contributing to the diagnostic process by capturing high-quality images and ensuring seamless collaboration with dermatologists.

## 6. Summary of the Evaluation

### 6.1. Planning

The requirements and criteria outlined in the usability plan following IEC 62366 are crucial for ensuring that the participants in the usability testing process are suitable and capable of providing valuable feedback on the eDerma platform. Here’s a summary of their importance:

**Requirements:** These are the essential criteria that all participants must meet to be eligible for the usability testing. They ensure that participants are of legal age, have the necessary equipment and connectivity, provide consent, and have the ability to understand instructions in English.

**Inclusion Criteria:** These criteria further refine the participant pool by ensuring that they are proficient in using the required technology, are willing to participate in the testing, and are part of the target user group (e.g., certified dermatologists, healthcare administrators, and health workers).

**Exclusion Criteria:** These criteria help exclude participants who do not meet the necessary qualifications or are unwilling to participate, ensuring that the testing group consists of individuals who can provide valuable insights into the usability and functionality of the eDerma platform.

Overall, these requirements and criteria are essential for ensuring the validity and reliability of the usability testing process, as they help select participants who are most representative of the platform’s intended users and can provide meaningful feedback to improve its usability and effectiveness.

### 6.2. Criteria Definition for each User Group

The criteria such as the requirements, inclusion and exclusion criteria for each user group is tabulated in the Table 5.

**Table 5:** Criteria Definition.

| Group | Requirement | Inclusion Criteria | Exclusion Criteria |
| --- | --- | --- | --- |
| **Patients** | 1. Shall be an adult population 18 years and older.<br>2. Shall have access to a computer or mobile device with internet connectivity.<br>3. Shall be able to read and understand instructions in English. | 1. Adult population 18 years and older.<br>2. Proficient in using a computer or mobile device.<br>3. Willing to participate in the usability testing. | 1. Below 18 years of age.<br>2. Inability to use a computer or mobile device proficiently.<br>3. Unwilling to participate in the usability testing. |
| **Dermatologists** | 1. Shall be a certified physician with the ability to diagnose and treat skin conditions.<br>2. Shall have access to a computer or mobile device with internet connectivity.<br>3. Shall have proficiency in using the eDerma platform. | 1. Certified physician.<br>2. Proficient in using the eDerma platform.<br>3. Willing to participate in the usability testing. | 1. Non-certified physician or students.<br>2. Inability to use the eDerma platform proficiently.<br>3. Unwilling to participate in the usability testing. |
| **Administrators** | 1. Shall have proficiency in healthcare software tools handling.<br>2. Shall have access to a computer or mobile device with internet connectivity.<br>3. Shall have proficiency in using the eDerma platform. | 1. Proficiency in healthcare software (EHR) tools handling<br>2. Proficient in using the eDerma platform.<br>3. Willing to participate in the usability testing. | 1. Healthcare professionals with no experience in healthcare software such as EHR<br>2. Inability to use the eDerma platform proficiently.<br>3. Unwilling to participate in the usability testing. |
| **Medical Photographers** | 1. Shall be a certified health worker who may have expertise in photography techniques.<br>2. Shall have access to a camera or mobile device with a camera.<br>3. Shall have proficiency in using the eDerma platform | 1. Certified health workers.<br>2. Shall have proficient in photography techniques.<br>3. Willing to participate in the usability testing. | 1. Non-certified health workers.<br>2. Inability to use the eDerma platform proficiently.<br>3. Unwilling to participate in the usability testing. |

These requirements and criteria are designed to ensure that the participants are suitable for the usability testing and can provide valuable feedback on the eDerma platform.

### 6.3. Setting

The usability study is done remotely, with the moderator interacting with the participants via Google Meet or Teams.

### 6.4. Preparation

Participants typically are sent the user manual before commencing the study with enough time to study it carefully. In some instances, as well as a review of the user manual, a training session may be required before users can access and interact with the software platform

### 6.5. Methods and Analysis

The feedback shall be gathered through a self-reported questionnaire and by direct observation of user behaviour. The questionnaire will consist of both quantitative and qualitative sections. In the quantitative section, users will rate various aspects of the software on a scale of 1 to 5, with 1 indicating the worst and 5 indicating the best. For the qualitative section, users will provide feedback on open-ended questions.

The quantitative data collected from the questionnaire will be used to calculate an average rating for each question. Based on these average ratings, decisions for further investigation will be made and documented in the usability evaluation report.

In addition, qualitative feedback will be analysed to assess whether the software is safe to use and if there are any unacceptable hazards associated with its usage. This qualitative analysis will also help determine if any modifications are needed to enhance the software’s safety and usability.

It’s worth noting that prior to the summative evaluation, a formative study was conducted to identify any initial usability issues and gather insights for refinement. The summative evaluation builds upon the findings of the formative study, providing a comprehensive assessment of the software’s usability and safety

## 7. Formative Study

The formative evaluation phase involved performing a usability study on three participants for each user group, totalling twelve sessions overall. During these sessions, users interacted with the software, completed assigned tasks, and provided feedback through self-reported questionnaires. Additionally, their behaviour and interactions with the software were observed and recorded.

Based on the feedback received and observations made during these sessions, adjustments were made to the usability questionnaires tailored for each user group. These modifications aimed to address any identified shortcomings and enhance the questionnaire’s effectiveness in gathering comprehensive feedback on the software’s usability.

The formative evaluation proved to be successful, as indicated by the feedback received from participants and the observed improvements in the effectiveness of the usability questionnaire. The insights gained from the formative study were invaluable in refining the questionnaires, ensuring their relevance and efficacy for the subsequent summative evaluation phase. The updated versions of the questionnaires, incorporating the refinements from the formative study, were then utilised for the summative evaluation process.

**Table 6:**
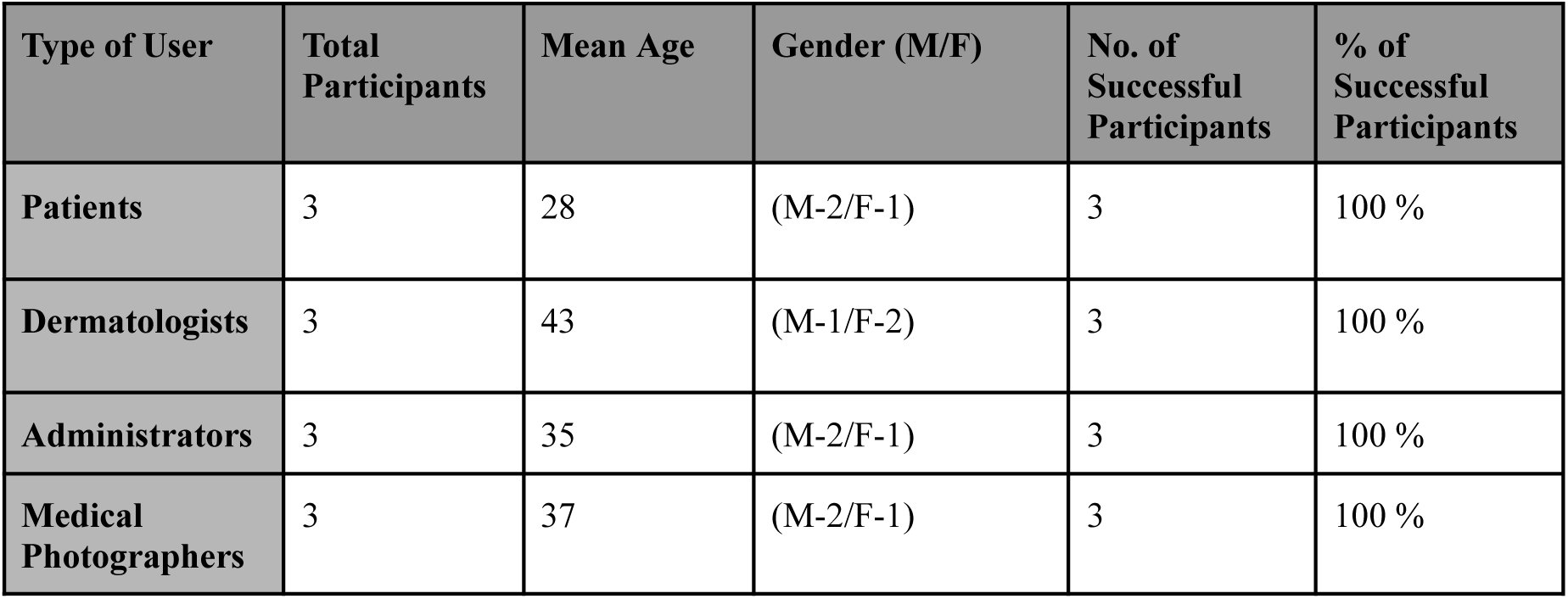
Formative Evaluation.

## 8. Summative Evaluation

The summative evaluation involved conducting usability assessments with a total of 12 participants for each user group, incorporating the insights gathered from the formative evaluation phase. These assessments aimed to comprehensively evaluate the usability of the software across diverse user groups, considering their unique perspectives and requirements.

Building upon the refinements made to the usability questionnaires based on the formative study findings, the summative evaluation sought to validate the effectiveness of the software in meeting user needs and expectations. Participants engaged with the software completed prescribed tasks, and provided feedback through structured questionnaires and open-ended discussions.

The streamlined approach, informed by the insights from the formative evaluation, ensured that the summative evaluation was conducted efficiently and effectively. Leveraging the lessons learned and adjustments made during the formative phase addressed potential usability issues proactively, contributing to a smoother evaluation process.

The results obtained from this phase, detailed in Section 9 of the article, provide valuable insights into the software’s usability strengths, areas for improvement, and overall effectiveness in meeting user needs. These findings serve as a foundation for further refinement and enhancement of the software, ultimately aiming to deliver an optimal user experience across all user groups.

**Table 7:** User Summative Evaluation.

| Type of User | Total Participants | Mean Age | Gender (M/F) | No. of Successful Participants | % of Successful Participants |
| --- | --- | --- | --- | --- | --- |
| Patients | 12 | 28 | (M-6/F-6) | 12 | 100 % |
| Dermatologists | 12 | 43 | (M-6/F-6) | 12 | 100 % |
| Administrators | 12 | 35 | (M-7/F-5) | 12 | 100 % |
| Medical Photographers | 12 | 37 | (M-5/F-7) | 12 | 100 % |

### 8.1 Use Errors and Hazards

The usability study identified no errors or hazards, and the participants could use the software correctly.

## 9. Results

The results section encompasses both quantitative and qualitative feedback obtained from participants during the usability evaluation of the Pathpoint® eDerma software. Quantitative feedback, detailed in Section 9.1, provides numerical ratings provided by participants for various aspects of the software’s usability. These ratings offer insights into participants’ perceptions of the software’s efficiency, effectiveness, and user-friendliness.

In Section 9.2, qualitative participant feedback delves into detailed open-ended responses and observations provided by participants. This qualitative feedback offers nuanced insights into participants’ experiences, preferences, and suggestions for improvement regarding the software’s usability.

Together, the quantitative and qualitative feedback paint a comprehensive picture of the software’s usability, highlighting strengths and improvement areas. By analysing both types of feedback, researchers can gain a deeper understanding of participants’ perspectives and use this information to refine and enhance the software further.

### 9.1. Quantitative Feedback from Participants

#### 9.1.1 Patients Group

The quantitative feedback from the patient group reflects high ratings across various aspects of the Pathpoint eDerma software’s usability. Participants provided ratings on a scale from 1 to 5 for different usability aspects, including User Interface Comfort, Responsiveness of the Platform, Clarity and User-friendliness, Interactive Experience, Quality, Accuracy, and Overall Usability rating.

Overall, participants rated the software favourably, with mean scores ranging from 4.57 to 5 across different usability aspects. Particularly noteworthy are the high mean ratings for Accuracy and Overall Usability, both receiving a perfect score of 5. This indicates that participants found the software to be highly accurate in its functionality and rated its overall usability very positively.

The highest mean rating was observed for Accuracy, with a perfect score of 5, indicating participants’ confidence in the software’s ability to deliver accurate results. Similarly, the Overall Usability rating received a perfect score of 5, reflecting participants’ overall satisfaction with the software’s usability. The mean performance of all participants in this user group, along with the two-time standard error of the mean against the corresponding features, is shown in Figure 1.

**Figure 1:**
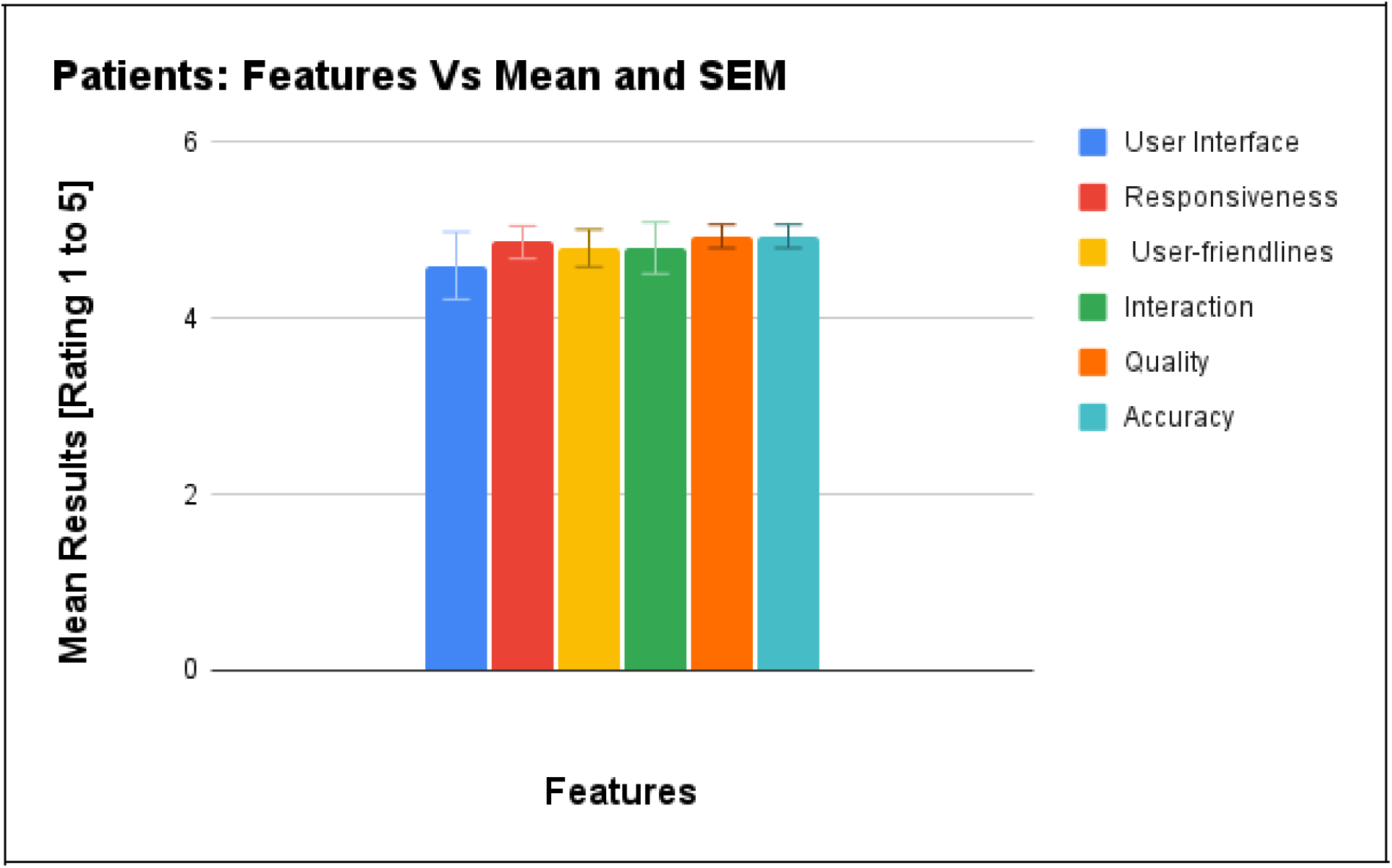
Performance of Patients User Group.

These quantitative results suggest that the Pathpoint eDerma software performed exceptionally well in meeting the usability needs and expectations of the patient group, as indicated by their high ratings across various usability aspects.

#### 9.1.2 Dermatologists Group

The quantitative feedback from the dermatologist’s user group indicates generally positive ratings for the usability of the Pathpoint eDerma software. Participants provided ratings on various usability aspects, including User Interface Comfort, Responsiveness of the Platform, Clarity and User-friendliness, Interactive Experience, Quality, Accuracy, and Overall Usability rating.

Overall, participants rated the software favourably, with mean scores ranging from 4.54 to 4.77. Notably, the highest mean rating was observed for Clarity and User-friendliness, with a score of 4.77, indicating participants’ satisfaction with the software’s clarity and ease of use. Similarly, the Responsiveness of the Platform received a mean rating of 4.62, suggesting participants found the platform adequately responsive to their needs.

The lowest mean rating was observed for User Interface Comfort, with a score of 4.54. However, this still indicates a generally positive perception of the software’s interface comfort among participants. Additionally, the Accuracy and Overall Usability rating received mean scores of 4.69, reflecting participants’ overall satisfaction with the software’s accuracy and usability. The mean performance of all participants in this user group, along with the two-time standard error of the mean against the corresponding features, is shown in Figure 2.

**Figure 2:**
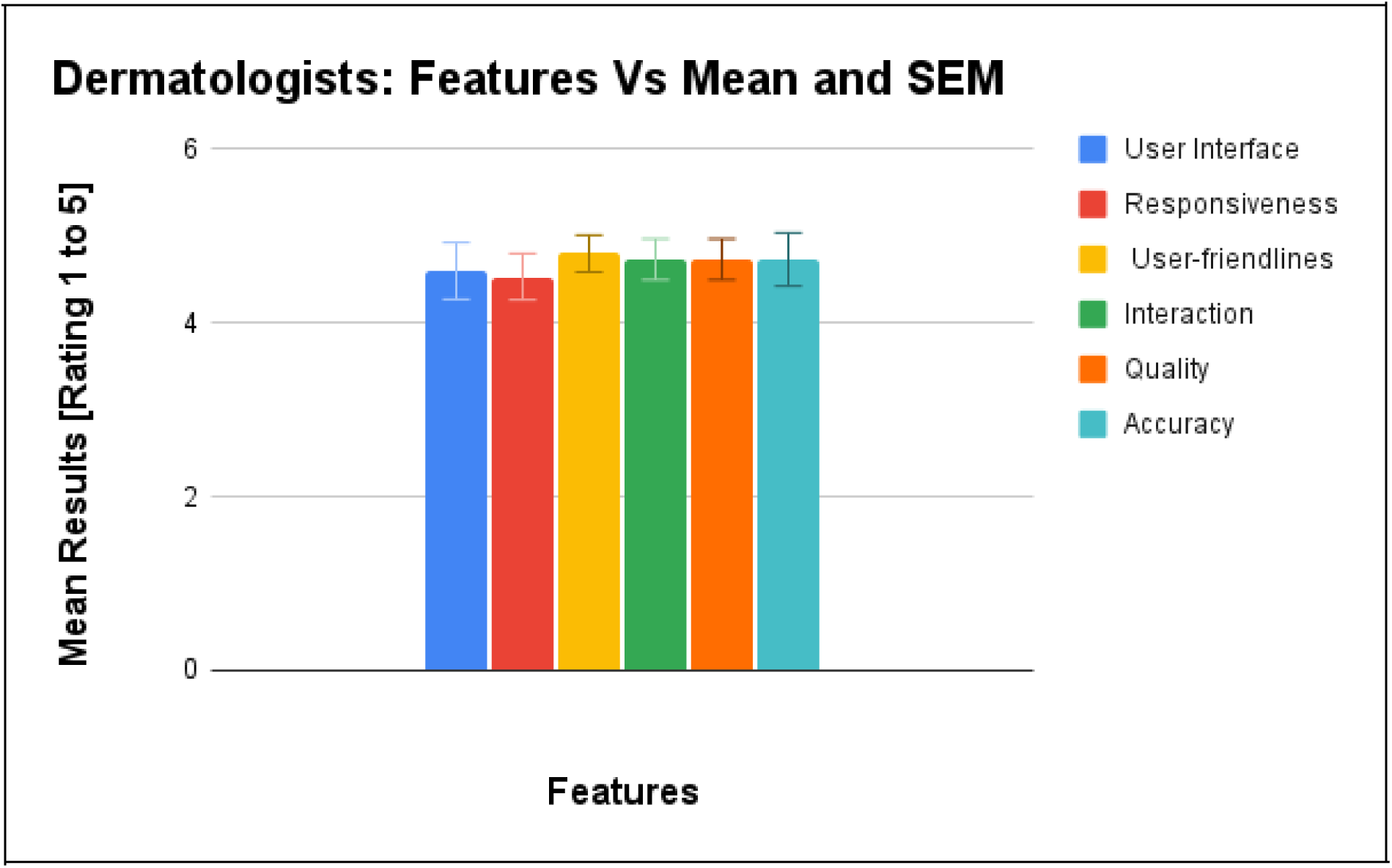
Performance of Dermatologists User Group.

These quantitative results suggest that while the Pathpoint eDerma software generally met the usability needs of the dermatologist’s user group, there may be some areas for improvement, particularly in enhancing interface comfort and addressing any issues related to accuracy.

#### 9.1.3 Administrators Group

The quantitative feedback from the administrators’ user group reveals positive ratings for the usability of the Pathpoint eDerma software across various dimensions. Participants provided ratings on key usability aspects, including User Interface Comfort, Responsiveness of the Platform, Clarity and User-friendliness, Interactive Experience, Quality, Accuracy, and Overall Usability rating.

Overall, participants rated the software favourably, with mean scores ranging from 4.33 to 4.87. Notably, the highest mean rating was observed for Interactive Experience, with a score of 4.93, indicating participants’ satisfaction with the software’s interactive features and user engagement. Similarly, Clarity and User-friendliness received a high mean rating of 4.87, suggesting that participants found the software intuitive and easy to use.

The lowest mean rating was observed for Responsiveness of the Platform, with a score of 4.33. However, this still indicates a generally positive perception of the software’s responsiveness among participants. Additionally, both Quality and Overall Usability ratings received mean scores of 4.80, reflecting participants’ overall satisfaction with the software’s quality and usability.The mean performance of all participants in this user group, along with the two-time standard error of the mean against the corresponding features, is shown in Figure 3.

**Figure 3:**
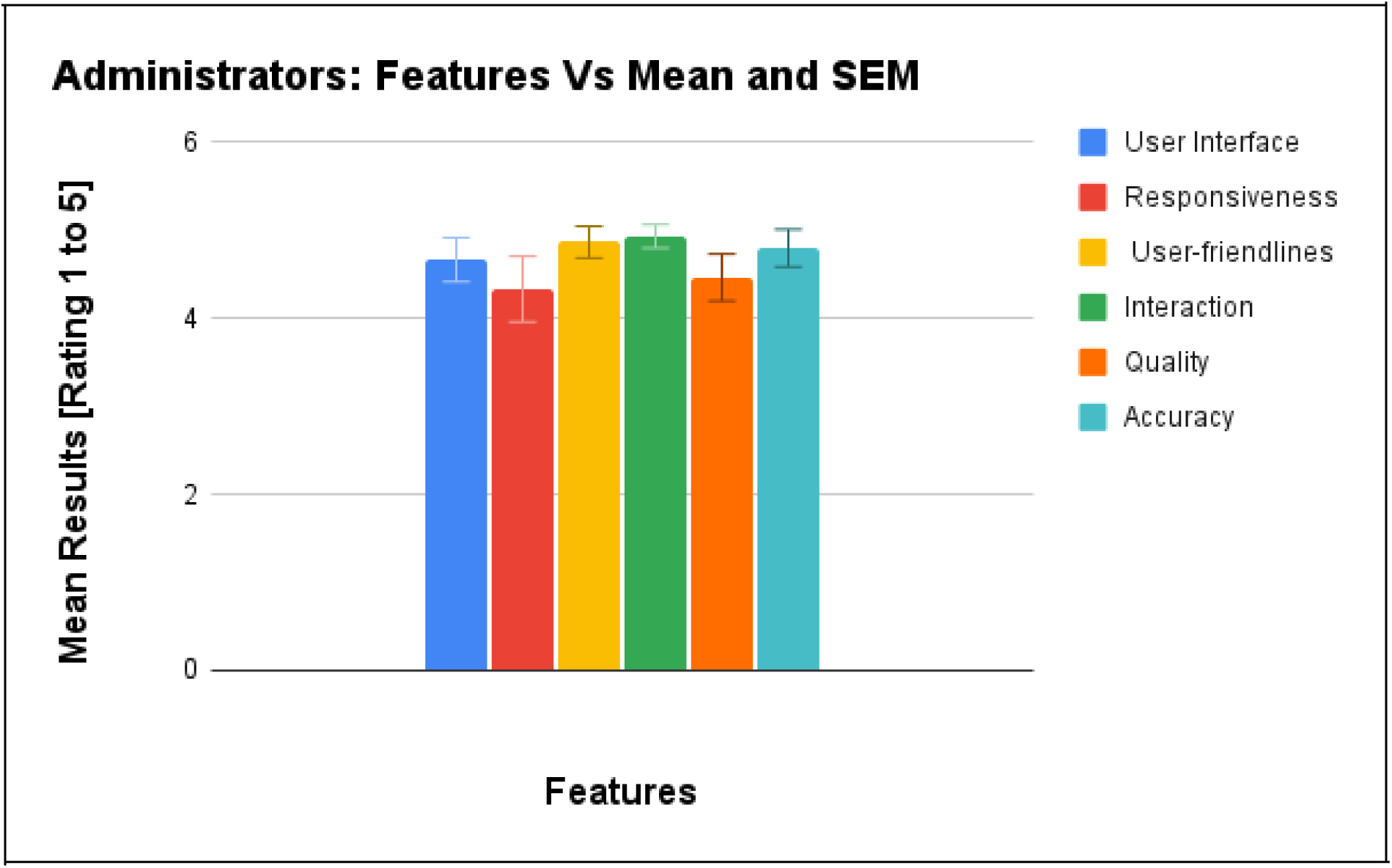
Performance of Administrators User Group.

These quantitative results suggest that while the Pathpoint eDerma software generally met the usability needs of the administrators’ user group, there may be some areas for improvement, particularly in enhancing platform responsiveness.

#### 9.1.4 Medical Photographers Group

The quantitative feedback from the medical photographer’s user group indicates overall positive ratings for the usability of the Pathpoint eDerma software. Participants provided ratings on various usability aspects, including User Interface Comfort, Responsiveness of the Platform, Clarity and User-friendliness, Interactive Experience, Quality, Accuracy, and Overall Usability rating.

Across all aspects, participants consistently rated the software highly, with mean scores ranging from 4.73 to 5. Notably, the Responsiveness of the Platform and Accuracy received perfect scores of 5, indicating participants’ satisfaction with the software’s responsiveness and accuracy in delivering results.

The highest mean rating was observed for Quality, with a score of 4.93, indicating participants’ high regard for the software’s quality in terms of performance and functionality. Similarly, the Overall Usability rating received a perfect score of 5, reflecting participants’ overall satisfaction with the software’s usability. The mean performance of all participants in this user group, along with the two-time standard error of the mean against the corresponding features, is shown in Figure 4.

**Figure 4:**
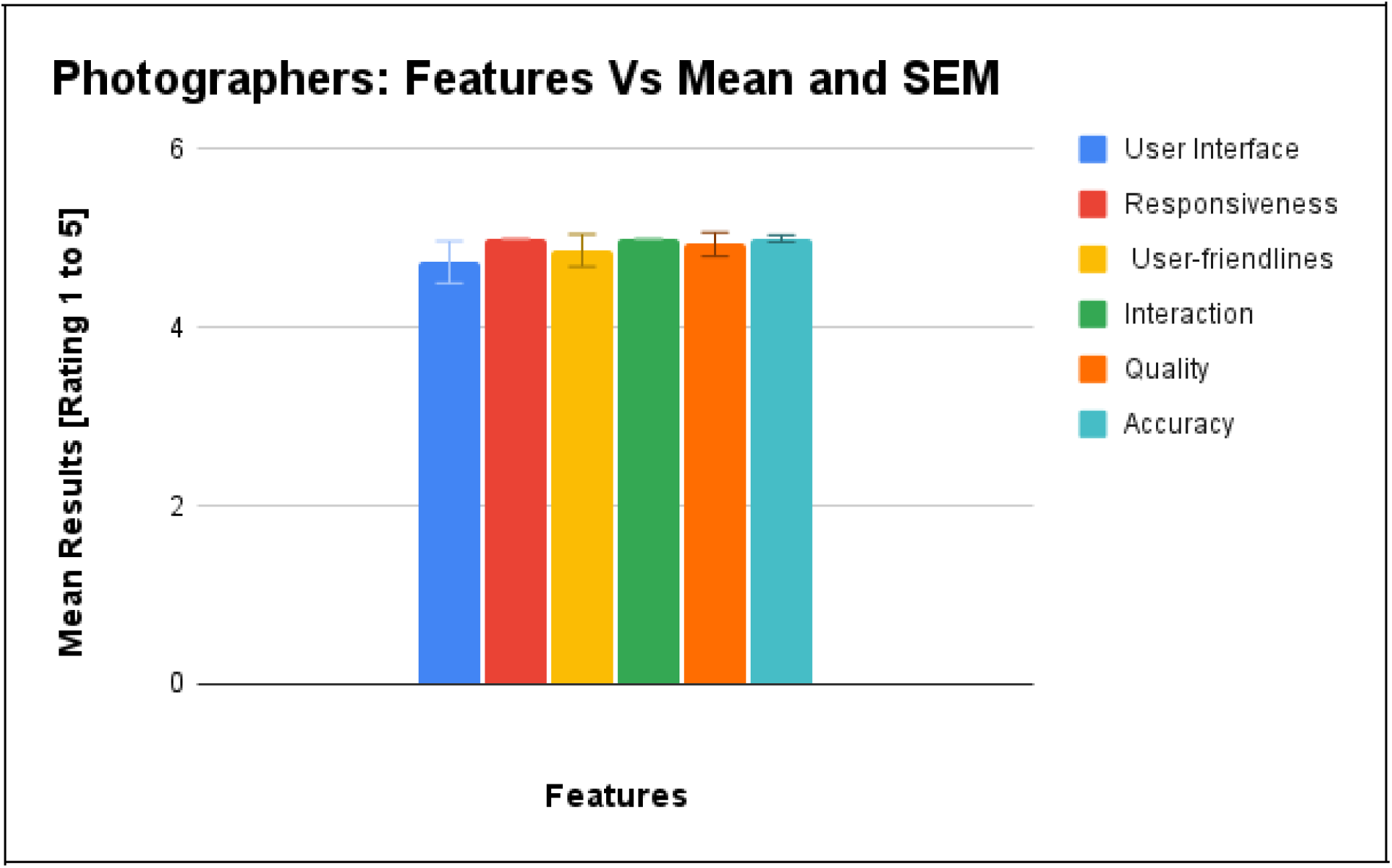
Performance of Medical Photographers User Group.

The overall performance of all the four user group, along with the two-time standard error of the mean against the corresponding features, is shown in Figure 5. These quantitative results suggest that the Pathpoint eDerma software effectively met the usability needs and expectations of the medical photographer’s user group, as indicated by their high ratings across various usability aspects.#

**Figure 5:**
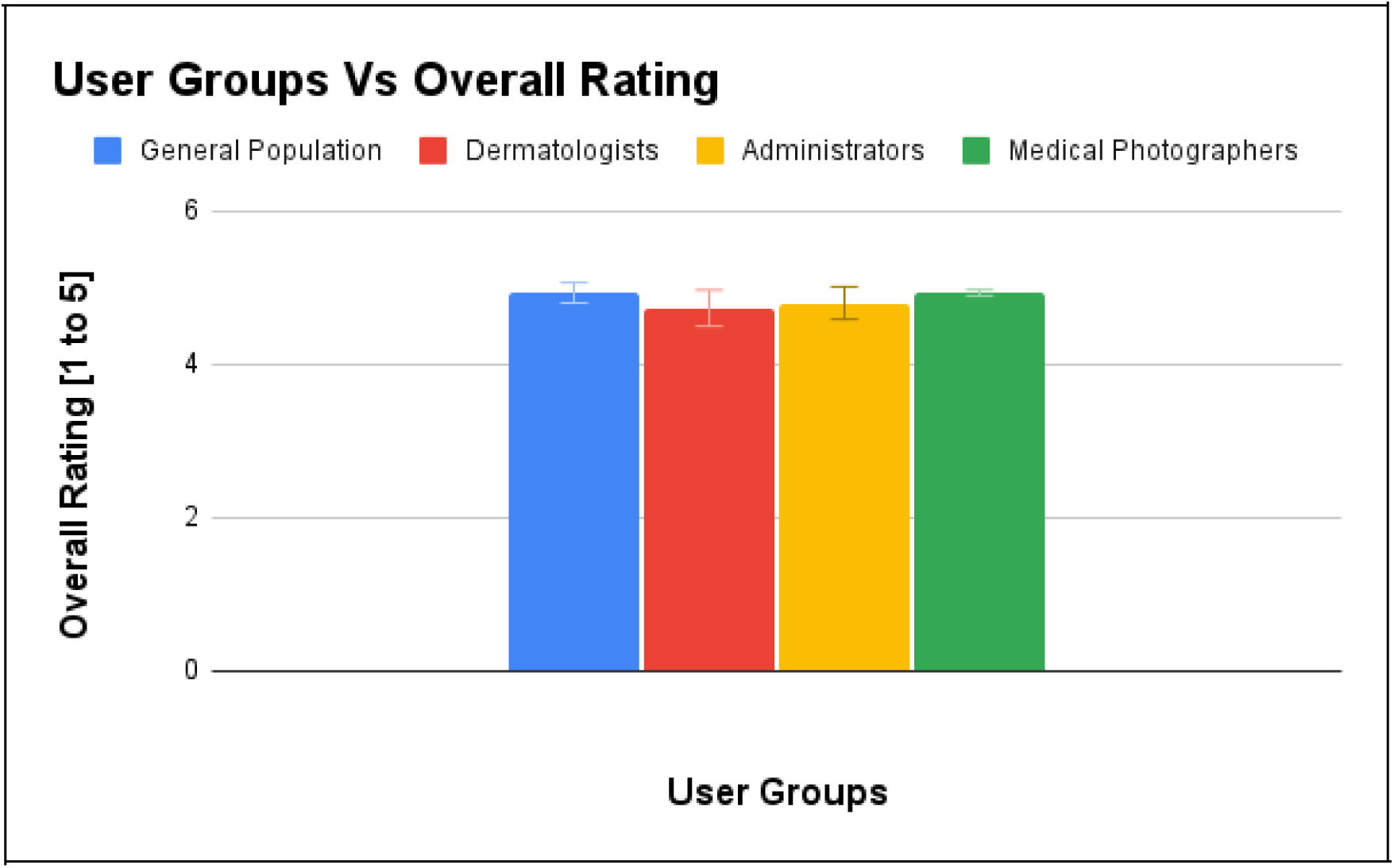
Performance Comparision of all User Group.

### 9.2. Qualitative Participant Feedback from Participants

Qualitative Participant Feedback serves as a rich source of user insights, offering a nuanced understanding of user experiences and perceptions during the usability evaluation of the Pathpoint® eDerma Software. This section captures participants’ voices, highlighting their observations, preferences, and suggestions that go beyond quantitative metrics. Through open-ended responses, users provide valuable context to their interactions with the software, shedding light on aspects such as user satisfaction, challenges faced, and recommendations for improvement. The qualitative feedback serves as a vital complement to quantitative data, offering a more holistic view of user sentiments and contributing to continuously enhancing the software’s usability and user experience

#### 9.2.1 Patients Group

##### Positive Feedback

- Users found it easy to locate and access the questionnaire in their emails, either through direct links or large buttons.
- The large buttons on the questionnaire contributed to a user-friendly experience, even on mobile devices.
- Navigation was straightforward, and the presence of videos in emails was deemed helpful.
- A significant number of users preferred remote assessments over face-to-face appointments, considering it a valuable and time-saving tool.

##### Areas for Improvement

- After completing the questionnaire, users suggested providing information about the next steps for better user engagement.
- The ‘Thank You’ page could be enhanced by displaying selected options to assure users that their responses were accurately recorded.
- Some users found the questionnaire wording lengthy, suggesting improvements in conciseness.
- Consideration for an option to increase font size for users with visual impairments was recommended.

#### 9.2.2 Dermatologists Group

##### Positive Feedback

- Users found the system user-friendly, straightforward, and easy to use, with a step-by-step flow facilitating patient action.
- The system was praised for its accessibility and comprehensive dashboard, providing all necessary information in one place for clinical decision-making.
- Users appreciated the ability to use the system remotely, which was seen as helpful in reducing waiting times and increasing efficiency.
- The platform was described as innovative and time-saving, allowing for quick assessment and diagnosis of patients.
- Communication between services was noted to be easy, contributing to a smoother workflow.
- Overall, users were impressed with the system’s user-friendliness, time-saving capabilities, and potential to improve patient dermatologist access.

##### Areas for Improvement

- Some users mentioned needing more practice with the system to provide more valuable feedback.
- Suggestions for improvement included adding more photos for each case and enhancing the font’s readability.
- While users generally found the system easy to use, there were minor suggestions for improvement, such as making the system capture all referrals and enhancing the templated letters.

#### 9.3.3 Administrators Group

##### Positive Feedback

- Users found the system user-friendly, straightforward, and easy to manage day-to-day tasks and workload.
- The system was described as clear, simple, and easy to understand quickly, with communication options such as comments and notifications being particularly useful.
- Users appreciated the design approach of “less is more” and liked the intuitive nature of the platform, which could potentially decrease waiting times.
- Different colours for buttons and tabs were praised for making it easy to differentiate and understand.
- The chronological presentation of patient lists and the clear buttons and timestamps labelling were helpful features.
- Users liked features such as tags and the ability to reduce waiting times, make services more accessible, and facilitate community and secondary care communication.

##### Areas for Improvement

- Users suggested improving the structure of the system to enhance usability.
- Differentiating between incoming and draft items could be improved visually.
- Some users mentioned the wordy assessment page and suggested simplifying it for better accessibility, especially for individuals with dyslexia.
- The delay in populating logs was identified as an area for improvement.

#### 9.3.4 Medical Photographers Group

##### Positive Feedback

- Users found the platform straightforward, easy to use, and similar to other platforms they’ve used before.
- They appreciated the clear buttons and the intuitive nature of the platform.
- The mandated PQ questions were liked, and users found it efficient to review images before saving.
- The platform was praised for cutting down waiting lists and facilitating virtual assessments, which benefits patients and medical photographers.
- The comment section was found to be helpful, and users liked that they could leave clinical comments if needed and visualise images after upload.
- The platform was described as self-explanatory, smooth, and not complex, resulting in time-saving benefits for medical photographers and clinical staff.
- Users appreciated features such as highlighting whether the PQ has been filled out and the automatic movement of referrals from the medical photographer’s list to the dermatologist’s list.

##### Areas for Improvement

- Users suggested a dark mode option, especially on phones, for better visibility.
- Some users mentioned the need for training and consideration of colour blindness and neurodiversity in the colour scheme.
- The visibility of the eye icon in the interface was identified as an area for improvement.

#### 9.3.5 Overall Feedback

Feedback from all user groups reflects a positive sentiment towards the system’s usability and functionality. Users commend its accessibility and efficiency, particularly in facilitating remote assessments and streamlining day-to-day tasks. However, there are consistent suggestions for improvement, such as enhancing communication clarity, providing customisation options for visual comfort, and refining labels for better intuitiveness. Addressing these areas could further optimise the system’s effectiveness and enhance the overall user experience.

## 10. Conclusion

The comprehensive usability evaluation conducted in accordance with established standards offers valuable insights into the system’s performance across diverse user groups. With a high success rate and no identified use errors or hazards, the evaluation underscores the software’s reliability and safety. Both quantitative metrics and qualitative feedback indicate positive user perceptions, particularly regarding accessibility, navigation, and overall functionality.

Users laud the user-friendly interface and efficiency in facilitating remote assessments while providing constructive suggestions for improvements in visual comfort and communication clarity. The absence of identified hazards reaffirms the platform’s reliability and suitability for real-world healthcare settings.

This evaluation is crucial in refining the system, ensuring it meets user expectations and aligns with industry standards. Actionable insights garnered from participants will guide continuous enhancements, ultimately delivering a safe, effective, and user-friendly platform for both general users and healthcare professionals. The study’s findings affirm the software’s usability characteristics and set the stage for successful deployment in healthcare environments.

## Data Availability

Data available on reasonable request

## 11. Acknowledgements

We want to express our gratitude to the participants for their valuable contributions to our SaMD usability study. Special thanks to Open Medical Ltd for their unwavering support, especially Mr. Jed Aubrey, throughout the research, which significantly enhanced the study’s success.

## References

ISO 62366-1:2015. (2015). “Medical devices—Part 1: Application of usability engineering to medical devices.” International Organization for Standardization.

Wiklund, M. E., & Kendler, J. (2018). “Usability Testing of Medical Devices.” CRC Press.

IEC/TR 62366-2:2016. (2016). “Medical devices—Part 2: Guidance on the application of usability engineering to medical devices.” International Electrotechnical Commission.

Karshmer, A. I., & Tenenberg, J. (2016). “Usability Evaluation in Industry.” CRC Press.

Bevan, N., & Macleod, M. (1994). “Usability Measurement in Context.” International Journal of Human-Computer Interaction, 6(1), 45–75.

Virzi, R. A. (1992). “Refining the Test Phase of Usability Evaluation: How Many Subjects is Enough?” Human Factors, 34(4), 457–468.

Nielsen, J. (1993). “Usability Engineering.” Academic Press.

Lewis, J. R. (1995). “IBM computer usability satisfaction questionnaires: Psychometric evaluation and instructions for use.” International Journal of Human-Computer Interaction, 7(1), 57–78.

Brooke, J. (1996). “SUS: A ‘Quick and Dirty’ Usability Scale.” In Usability Evaluation in Industry (pp. 189–194). CRC Press.

Dumas, J. S., & Redish, J. C. (1999). “A Practical Guide to Usability Testing.” Intellect Books.

Shneiderman, B. (1998). “Designing the User Interface: Strategies for Effective Human-Computer Interaction.” Addison-Wesley.

Kushniruk, A. W., & Patel, V. L. (2004). “Cognitive and usability engineering methods for the evaluation of clinical information systems.” Journal of Biomedical Informatics, 37(1), 56–76.

Marcilly, R., Ammenwerth, E., & Roehrer, E. (2006). “Assessing Usability of User Interfaces for Clinical Information Systems: Which Framework(s) Should Be Used?” Stud Health Technol Inform, 124, 343–348.

Rubin, J., & Chisnell, D. (2008). “Handbook of Usability Testing: How to Plan, Design, and Conduct Effective Tests.” Wiley.

Patel, M. R., Kegelmeyer, D. A., & Kegelmeyer, W. P. (2017). Usability testing in the context of IEC 62366. In 2017 IEEE International Conference on Healthcare Informatics (ICHI) (pp. 505-509). IEEE.

Hignett, S., Lang, A., Pickup, L., & McLeod, R. (2018). Applying IEC 62366:2015 in usability engineering. Applied Ergonomics, 68, 187–195.

Nielsen, C., Overgaard, M., & Pedersen, M. (2019). Integrating IEC 62366 in Usability Studies - A Case Study. In International Conference on Human-Computer Interaction (pp. 335-349). Springer.

Smith, S. N., Mosleh, A., & Jiang, X. (2020). A systematic review of usability evaluation methods for medical devices. BMC Medical Informatics and Decision Making, 20(1), 1–19.

Balamurugan Subramaniyan, Piyush Mahapatra, Muna Mohamud, & Atlas Naqvi (2024). Comprehensive Usability Evaluation and Performance Overview of Pathpoint Outcomes Software. doi: 10.1101/2024.05.11.24307045. medRxiv Preprint.

